# Geo-Demographic Trends in Nontraumatic Subarachnoid Hemorrhage-Related Mortality Among Older Adults in the United States, 1999-2020

**DOI:** 10.1101/2023.12.25.23300527

**Authors:** Martin G. McCandless, Anand A. Dharia, Elizabeth E. Wicks, Paul J. Camarata

## Abstract

**Introduction:** Nontraumatic subarachnoid hemorrhage (ntSAH) often results from a ruptured aneurysm and correlates with significant morbidity and mortality, particularly among the older population. Despite its impact, limited comprehensive studies evaluate the longitudinal trends in ntSAH-related mortality in older adults in the United States (US).

**Methods:** The authors conducted a retrospective analysis using the CDC WONDER database from 1999 to 2020, analyzing Multiple Cause-of-Death Public Use death certificates to identify ntSAH as a contributing factor in the death of adults aged 65 years and older. We calculated age-adjusted mortality rates (AAMR) and annual percent change (APC) to examine trends across demographic variables such as sex, race/ethnicity, urbanization, and states/census region.

**Results:** 78 260 ntSAH-related deaths (AAMR 8.50 per 100 000 individuals) occurred among older adults in the US from 1999 to 2020. The overall AAMR for ntSAH decreased from 9.98 in 1999 to 8.67 in 2020 with an APC of −0.7% (95% CI [−1.0, −0.3]). However, the authors observed a noticeable rise from 2013 to 2020 with an APC of 1.7% (95% CI [0.8, 2.6]). Sex, racial, and regional disparities were evident with higher mortality rates for ages 85 or greater (crude mortality rate 16.6), women (AAMR 9.55), non-Hispanic Asian or Pacific Islander (AAMR 12.5), and micropolitan areas (AAMR 8.99), and Western US (AAMR 8.65).

**Conclusion:** Mortality from ntSAH increases with age, affects women disproportionately, and occurs more often in an inpatient setting. These findings necessitate targeted, multi-dimensional health policies and clinical interventions. Specialties beyond neurosurgery can utilize this data for improved risk stratification and early treatment. Policymakers should focus on equitable resource allocation and community-level interventions to mitigate these trends effectively.

## Introduction

The risk of nontraumatic subarachnoid hemorrhage (ntSAH) rises sharply with age.^1–3^ Due to aging demographics in the United States (US), ntSAH warrants a more focused epidemiological evaluation by American neurosurgeons. Especially among adults aged 65 years and older, the morbidity and mortality associated with ntSAH represent a considerable clinical and economic burden.^4^

A growing body of literature suggests advancements in diagnostic modalities and treatment interventions have improved the overall outcomes of SAH patients.^5^ But have these advancements equally benefited older adults? Age-specific data on ntSAH-related mortality among the older population in the US remains minimal.^6^ The lack of comprehensive analysis could impede the formation of targeted health policy measures aimed at reducing mortality and improving the quality of life in this vulnerable group.^7^

Given the demographic shifts and the aging population’s susceptibility to ntSAH, we investigate how trends in ntSAH-related mortality continue to evolve among adults aged 65 and above. To address this question, this study performs an in-depth analysis of data from the Centers for Disease Control and Prevention’s Wide-ranging Online Data for Epidemiologic Research (CDC WONDER) database. By dissecting these trends, we aim to contribute to existing knowledge on ntSAH epidemiology, thereby informing clinical guidelines and health policy initiatives to mitigate the impact of ntSAH-related mortality among the older population.

## Methods

### Data Source

This study investigated the number of deaths associated with nontraumatic subarachnoid hemorrhages (ntSAH) from ages 65 years and older in the United States from 1999 to 2020. The study used the CDC WONDER database.^8^ This study was exempt from institutional review board approval due to the public and anonymous nature of the database and was compliant with STROBE guidelines.

This study analyzed Multiple Cause-of-Death Public Use death certificates to identify ntSAH as a contributing factor in the death. The authors obtained the number of annual deaths related to ntSAH and the corresponding population sizes. The data were separated into categories based on sex, race/ethnicity, ten-year age groups, level of urbanization, and state. The study used the *International Statistical Classifications of Diseases, Tenth Revision* (ICD-10) code for ntSAH (I60.x).^9^

The sex, ethnicity, and race were recorded on the death certificates in compliance with the US Office of Management and Budget.^10^ Ethnicity and race were divided into separate categories, including Hispanic or Latino, non-Hispanic Black or African American, non-Hispanic Asian or Pacific Islander, or non-Hispanic White. Although we included the individuals who did not fall into the categories of Hispanic or Latino or non-Hispanic in the overall analysis, we omitted these from the subgroup analysis due to the suppression of data with too few individuals in CDC WONDER.

Urbanization was defined in compliance with the 2013 National Center for Health Statistics Urban-Rural Classification Scheme for Counties.^11^ Metropolitan was subdivided into large central metros (population > 1 million and completely contain or are entirely contained by the largest city or contain at least 250 000 residents of any city within the statistical area), large fringe metros (population > 1 million and not central metro), medium metros (population 250 000 – 999 999), and small metros (population < 250 000). Nonmetropolitan counties are subdivided into micropolitan (population 10 000 – 49 999) and noncore (population < 10 000). The data for each state excluded the District of Columbia. The place of death was subdivided into inpatient, nursing home, home, hospice facility, and other/unknown.

### Statistical Analysis

The crude mortality rate (CMR) and age-adjusted mortality rates per 100 000 (AAMR) were calculated using the US 2000 standard population.^12^ To evaluate mortality trends, Joinpoint Regression Program (v4.9.1.0; National Cancer Institute, 2022)^13^ was employed to analyze the overall population and subgroup segments. The Monte Carlo permutation test was employed to determine the annual percent change (APC) in mortality with 95% confidence intervals at identified segments linking join points. The slopes of APCs were then calculated and assessed using a two-sided t-test against a null value of 0. Statistical significance was determined by a P-value < 0.05. Analysis was conducted in October 2023.

## Results

A total of 78 260 ntSAH-related deaths (AAMR 8.50 per 100 000 individuals) occurred among older adults in the United States from 1999 to 2020. Of the total deaths, 26 285 (33.9%) were men, 51 359 (66.1%) were women. Of the known ethnic and racial groups 60 151 (76.9%) were non-Hispanic White, 6 873 (8.8%) were non-Hispanic Black or African American, 4 118 (5.3%) were non-Hispanic Asian or Pacific Islander, and 5 963 (7.6%) were Hispanic or Latino. Of the 75 928 known locations of death, 59 122 (75.6%) were inpatient facilities, 8 004 (10.2%) were nursing homes, 4 911 (6.3%) were in the decedent’s home, and 3 891 (5.0%) were in hospice facilities. Among diagnosis, age, sex, race and ethnicity, and urbanization, the highest mortality rates were seen in those 85 or older (CMR 16.9), women (AAMR 9.55), non-Hispanic Asian or Pacific Islander adults (AAMR 12.5), micropolitan populations (AAMR 8.99), and in the Western US (AAMR 8.65) (Table 1, eTable 1).

**Table 1.** Demographics of Nontraumatic Subarachnoid Hemorrhage-Related Mortality.

| Characteristics | Deaths (%)<sup>1</sup> | CMR or AAMR [95% CI] |
| --- | --- | --- |
| Total | 78,260 (100) | AAMR: 8.50 [8.44, 8.56] |
| Ten-Year Age Groups |  | CMR |
| 65-74 Years | 27,905 (35.9) | 5.47 [5.40, 5.53] |
| 75-84 Years | 29,591 (38.1) | 9.91 [9.80, 10.0] |
| 85+ Years | 20,148 (26.0) | 16.9 [16.6, 17.1] |
| Gender |  | AAMR |
| Men | 26,285 (33.9) | 6.94 [6.86, 7.03] |
| Women | 51,359 (66.1) | 9.55 [9.47, 9.63] |
| Race / Ethnicity |  | AAMR |
| Non-Hispanic White | 60,151 (76.9) | 8.06 [8.00, 8.13] |
| Non-Hispanic Black or African American | 6,873 (8.78) | 8.87 [8.66, 9.08] |
| Hispanic or Latino | 5,963 (7.62) | 9.67 [9.42, 9.92] |
| Non-Hispanic Asian or Pacific Islander | 4,118 (5.26) | 12.5 [12.2, 12.9] |
| Non-Hispanic Native American or Alaska Native | 385 (0.49) | 8.73 [7.83, 9.62] |
| Other/unknown | 770 (0.98) |  |
| Urbanization |  | AAMR |
| Noncore | 6,071 (7.81) | 8.19 [7.99, 8.40] |
| Micropolitan | 8,259 (10.6) | 8.99 [8.80, 9.19] |
| Small Metro | 7,889 (10.2) | 8.53 [8.34, 8.71] |
| Medium Metro | 16,978 (21.9) | 8.59 [8.46, 8.72] |
| Large Fringe Metro | 17,428 (22.5) | 8.03 [7.91, 8.15] |
| Large Central Metro | 21,019 (27.1) | 8.46 [8.35, 8.58] |
| Census Region |  | AMMR |
| Northeast | 15,209 (19.4) | 8.34 [8.21, 8.48] |
| Midwest | 17,768 (22.7) | 8.54 [8.41, 8.66] |
| South | 27,799 (35.5) | 8.32 [8.22, 8.42] |
| West | 16,868 (21.6) | 8.65 [8.51, 8.78] |
| Unknown | 616 (0.79) | - |
| Location of Death |  |  |
| Inpatient | 59,122 (75.6) | - |
| Nursing Home | 8,004 (10.2) | - |
| Home | 4,911 (6.28) | - |
| Hospice Facility | 3,891 (4.97) | - |
| Other/Unknown | 2,332 (2.98) | - |

<sup>1</sup>Percentage may not sum to 100 due to rounding. CMR: Crude mortality rate per 100,000 individuals; AAMR: Age-adjusted mortality rate per 100,000 individuals

The overall AAMR for ntSAH in older adults decreased from 9.98 in 1999 to 8.67 in 2020 with an average annual percent change (AAPC) of −0.7%, 95% CI [−1.0, −0.3]. The AAMR decreased from from 1999 to 2013 (APC, −1.8% 95% CI [−2.2, −1.5]) followed by a rise from 2013 to 2020 (APC, 1.7% 95% CI [0.8, 2.6]). NtSAH-related mortality remained stable for men but decreased for women. Specifically, the AAMR in men changed from 7.40 in 1999 to 7.89 in 2020, while the AAMR for women decreased from 11.7 in 1999 to 9.27 in 2020. Although the AAMR for both men and women decreased from 1999 to 2014 and 1999 to 2011, respectively, the AAMR for men increased from 2014 to 2020 (APC, 2.9% 95% CI [1.7, 4.2]) while the AAMR for women had a statistically insignificant rise from 2011 to 2020 (APC, 0.6% 95% CI [−0.1, 1.3]). (Figure 1)

**Figure 1.**
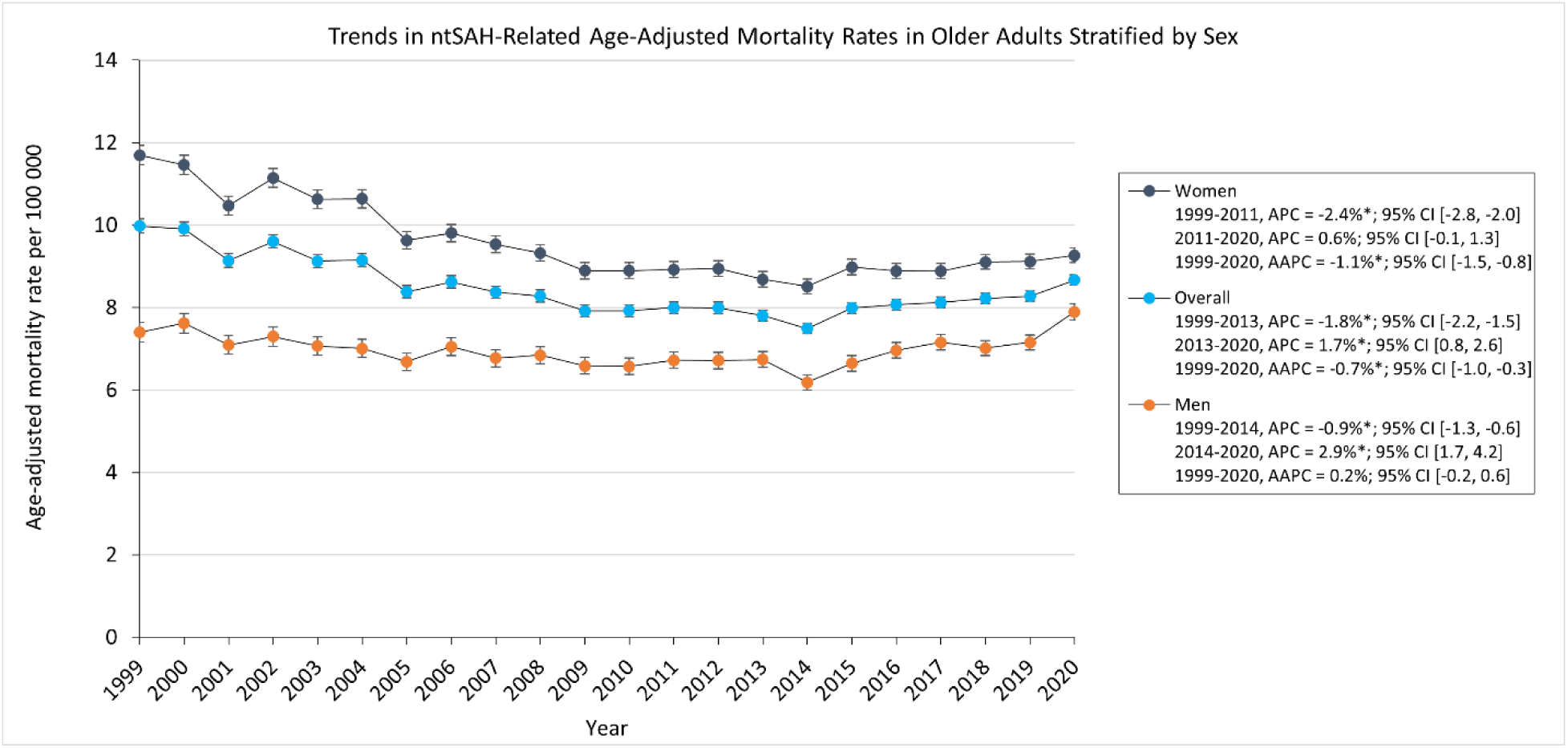
Trends in ntSAH-Related Age-Adjusted Mortality Rates in Older Adults Stratified by Sex.

AAMR decreased for all ethnic and racial groups. Asian or Pacific Islander adults had the highest AAMR which decreased from 17.2 in 1999 to 12.0 in 2020. The AAMR for White, Black, and Asian or Pacific Islander adults decreased from 1999 to 2012, 2013, and 2014, respectively, followed by a statistically significant increase to 2020 for White and Black adults. The AAMR for Asian or Pacific Islander adults had a statistically insignificant increase (APC, 1.1% 95% CI [−1.4, 3.7]), while the AAMR for Hispanic or Latino adults decreased from 13.7 in 1999 to 9.30 in 2020. (Figure 2)

**Figure 2.**
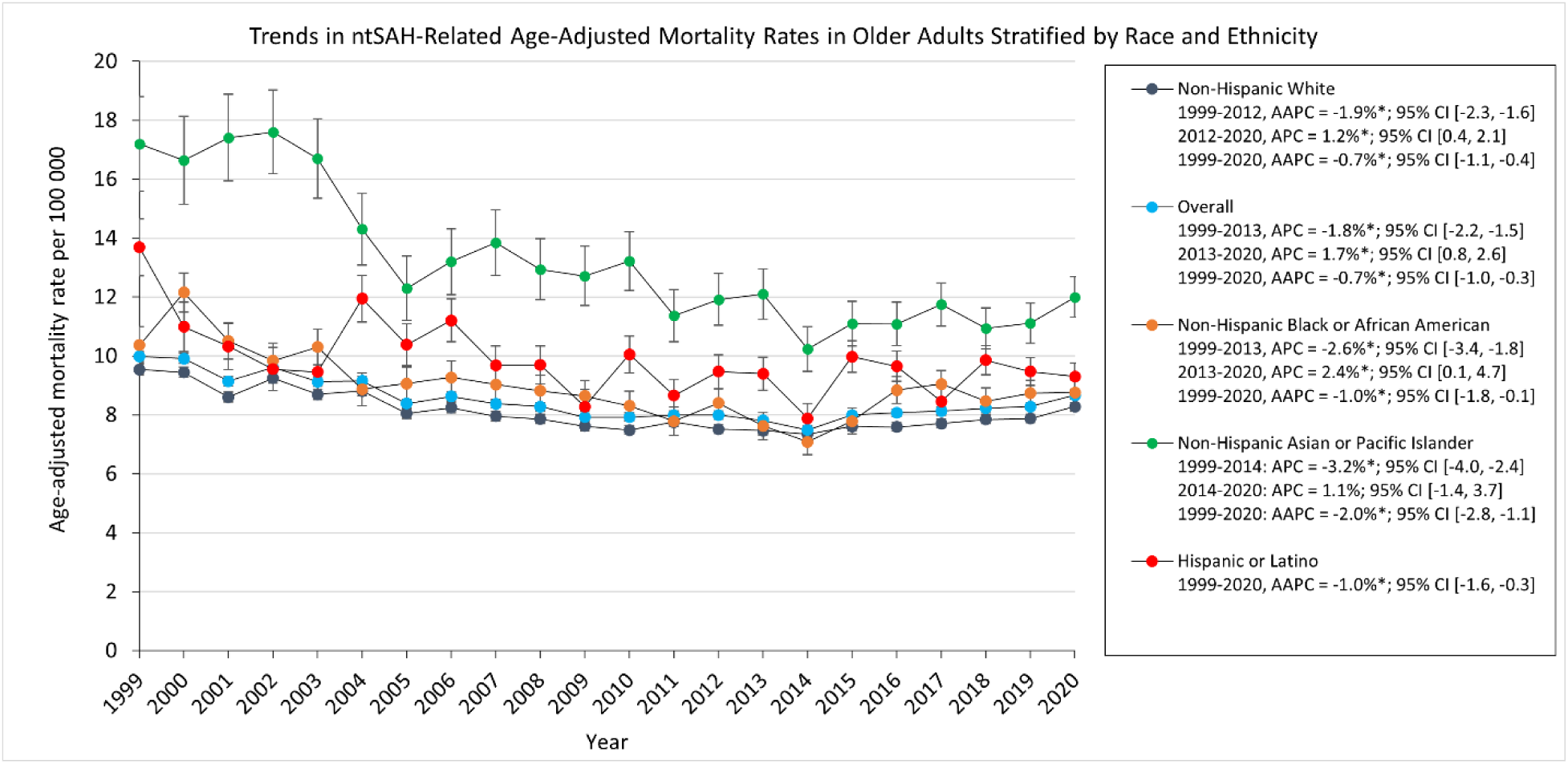
Trends in ntSAH-Related Age-Adjusted Mortality Rates in Older Adults Stratified by Race and Ethnicity.

Subgroup analysis showed that CMR in those 65 and older decreased from 9.93 in 1999 to 8.32 in 2020. The CMR for adults 65-74 and 75-84 years old decreased from 1999 through 2020 while the CMR in adults 85 or older increased from 16.0 in 1999 to 19.3 in 2020 (eFigure 1). From 1999 through 2020, the proportions of ntSAH-related deaths decreased in inpatient facilities (86.0% to 66.7%). In contrast, the proportion of ntSAH-related deaths increased in homes (3.7% to 11.4%), nursing homes (9.3% to 9.9%), and hospice facilities (0.4% in 2003 to 9.6% in 2020) (eTable 2, eFigure 2). The AAMR of ntSAH as the underlying cause of death decreased from 1999 to 2013 (APC, −2.5% 95% CI [−2.7, −2.2]) before stabilizing from 2013 to 2020 (APC, 0.2% 95% CI [−0.4, 0.9]) (eTable 3, eFigure 3)

Mortality rates for all urbanization groups decreased from 1999 to 2020 except for micropolitan and noncore areas which remained stable. Micropolitan areas had the highest AAMR which decreased from 11.2 in 1999 to 9.93 in 2020. All urbanization groups decreased from 1999 to the 2010s. The AAMR for noncore, micropolitan, and medium metropolitan areas increased from 2008, 2011, and 2012 through 2020, respectively, while medium metropolitan, large fringe and large central areas remained stabilized from 2012 and 2014 through 2020, respectively. (Figure 3).

**Figure 3.**
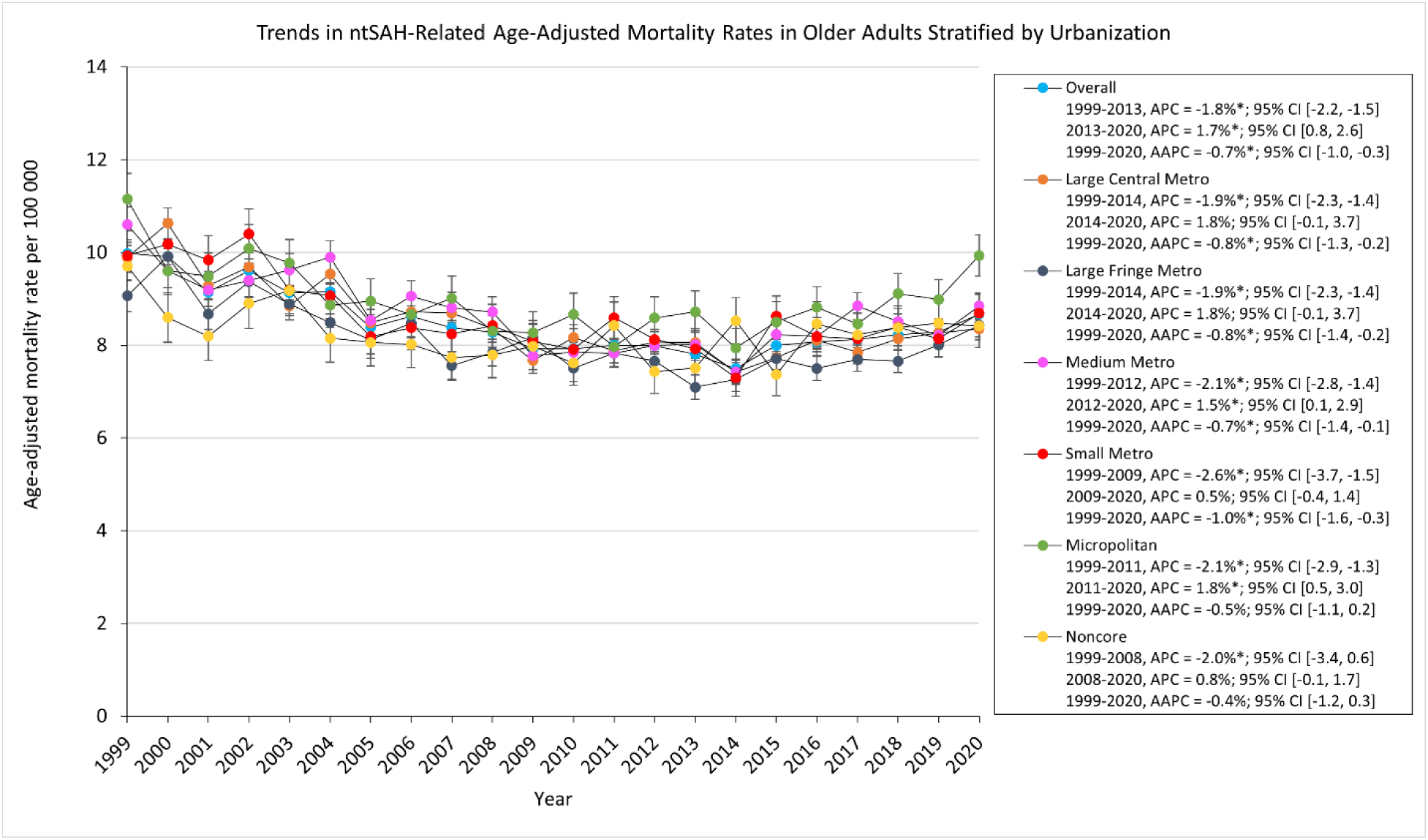
Trends in ntSAH-Related Age-Adjusted Mortality Rates in Older Adults Stratified by Urbanization.

There were significant regional differences in the burden of ntSAH-related mortality with the Western US having a significantly higher mortality burden compared to the remaining census regions. (Table 1, eTable 4). The top 5 states (Hawaii, Tennessee, West Virginia, Alaska, and Washington) with the highest ntSAH-related mortality had much higher mortality than those in the bottom 5 states (Kansas, New Hampshire, Arizona, Florida, and Maine) (Figure 4).

**Figure 4.**
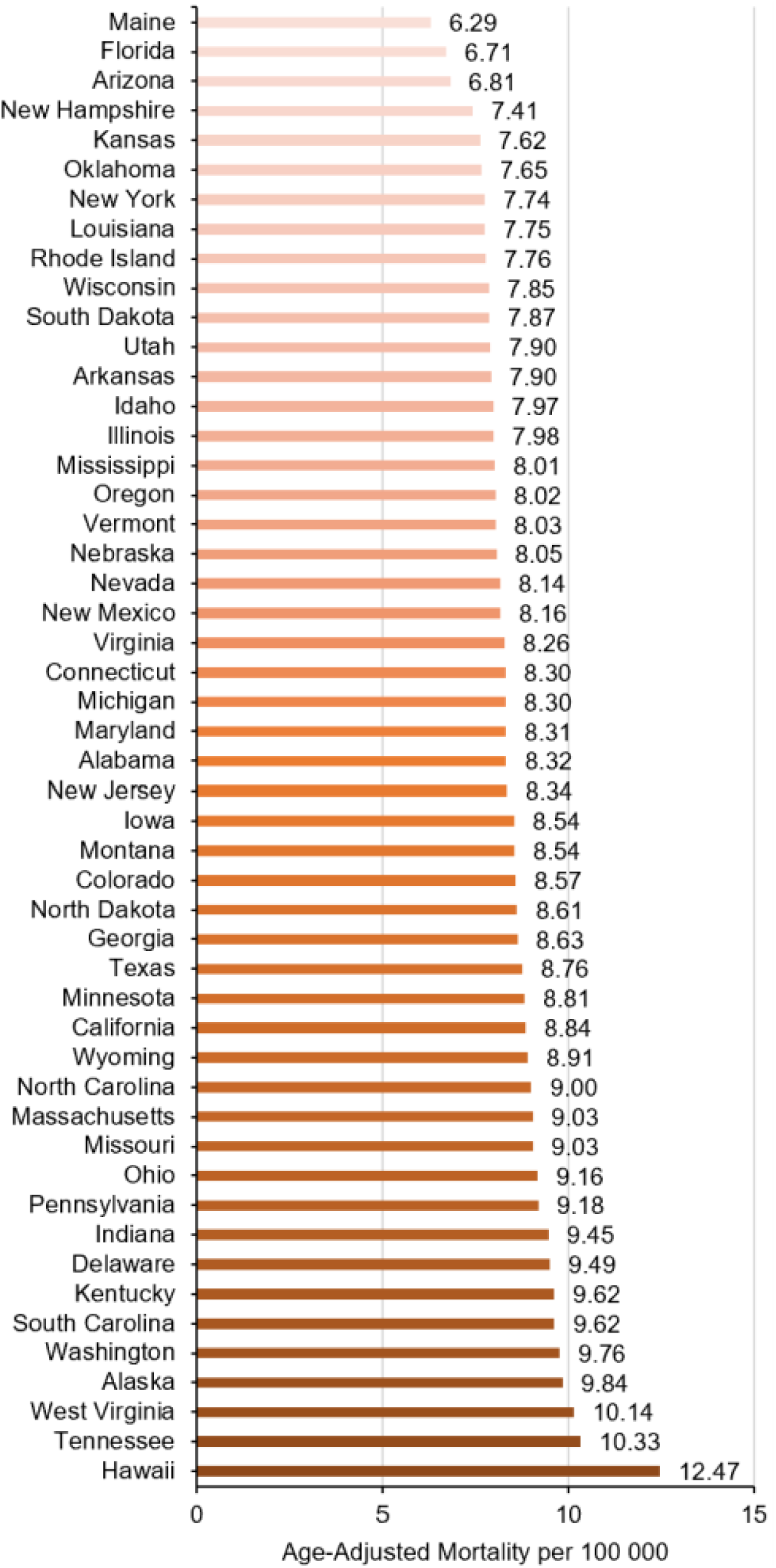
Trends in ntSAH-Related Age-Adjusted Mortality Rates in Older Adults Stratified by State.

## Discussion

This comprehensive analysis of ntSAH-related mortality among adults aged 65 years and older from the CDC WONDER database from 1999 to 2020 demonstrates a complex trajectory. While the overall AAMR exhibited an annual decline of −0.7%, the overarching figure conceals dynamic changes over the span of 2 decades. An initial decline in mortality was observed from 1999 to 2013, followed by a concerning rise from 2013 to 2020. This resurgence in mortality rates, despite advancements in neurosurgical interventions and medical management, signals a need for further investigation into evolving risk factors and healthcare disparities in this vulnerable population.

ntSAH is a potentially fatal disease that traditionally affected middle-aged patients younger than the age of 60 years; however, with improvements in overall health care and longer life expectancies, the incidence of elderly patients with ntSAH is increasing.^14–16^ NtSAH predominantly refers to intracranial bleeding within the subarachnoid space from a cerebral aneurysm rupture. Although only an estimated 15-20% of patients with ntSAH are greater than 70 years old, elderly patients incur relatively high total healthcare expenditures, long-term disability, and mortality.

As patient demographics have shifted towards an older age, clinical practice has also evolved with more elderly ntSAH patients referred to neurosurgical centers than ever before. Nearly 2 decades after the publication of the International Subarachnoid Aneurysm Trial (ISAT), which highlighted the utility of endovascular interventions over traditional microsurgical clipping and craniotomy for ruptured cerebral aneurysms, the treatment of ntSAH has changed dramatically.^17^ In support of aggressive management of cerebral aneurysms and accommodation of elderly patients unable to tolerate surgery, the ISAT results have led to the explosive growth of endovascular coiling in the elderly SAH population. Although increasing technological sophistication has led to increased device costs and healthcare expenditures, we are now better equipped to treat patients with ntSAH.^18^

Nevertheless, the appropriate treatment of ntSAH in elderly patients, particularly for occlusion of ruptured intracranial aneurysms, is controversial and challenging. Rebleeding is the most feared complication after SAH, which may still result in death or permanent dependence in 80% of instances.^19^ If left untreated, aneurysms may rebleed in the first month after a hemorrhage in 40% of cases. On the other hand, treatment of aneurysms itself, either by operative or endovascular occlusion, portends significant inherent risk and must be weighed in the overall clinical context. Even those undergoing successful treatment, many suffer delayed cerebral ischemic events or other systemic complications.

Several studies have cited advancing age, along with the severity of clinical presentation on admission, as two of the strongest prognostic factors for predicting poor functional outcomes.^20–23^ Age negatively impacts survival and recovery as older patients have a greater number of co-morbidities and systemic complications. Moreover, ntSAH has a unique pathophysiology in elderly patients. Elderly patients have greater degrees of brain atrophy, vessel calcification or stenosis, and hypertension, which may affect the pattern and extension of blood after a rupture. Additionally, elderly patients have lower cerebral blood flow and lower cardiac output. While less elasticity of the vessels may lower the probability of vasospasm after ntSAH, elderly patients have a high likelihood of mortality and cardiovascular events compared with the younger healthy population.^24,25^ Although these age-related pathophysiologic differences may account for greater mortality in elderly patients, the increasing mortality rate since 2013 within the cohort is alarming. A greater number of treatments, especially endovascular interventions enhanced by rapidly advancing technologies, are now offered to elderly patient more than at any previous time. The authors suspect that with improved access to tertiary care centers and greater number of elderly patients considered for treatment, more deaths are now coded appropriately as ntSAH. This is compounded by the number of elderly individuals or family members who may choose to withdraw care and clinicians who forgo treatment understanding the poor functional outcomes within this cohort.

Which patients are considered elderly? Identifying at-risk populations with strict definitions can be important for developing risk stratification measures to optimize timely treatment and improve patient outcomes. In this study, older patients were defined as those above the age of 65 years, as stratified by the CDC WONDER database; however, the cutoff for elderly patients is not strict within the literature. The definition of elderly patients has changed from 60 and up to 75-80 years and older. Age cut-offs of 65, 75, and 80 years are the most common variations reported in studies that potentiate differences in treatment paradigms. While many authors recommend aggressive aneurysm treatment for patients admitted in good clinical condition, others factor in advanced age. Goldberg et al. showed that the decision to treat or pursue conservative management differed significantly for patients aged 80 to 90 years, leading to a possible overestimation of mortality and unfavorable outcomes in this group.^26^ This could also reflect less interventions desired at the patients’ request. Generalization by aggregating patients above the age of 65 may limit the ability to make targeted reforms.

Mortality rates for ntSAH showed substantial sex-based disparities. Our studies corroborate the findings of Xia, which showed persistently higher mortality from SAH in women aged 65 years and older.^27^ This greater mortality reflects the higher percentage of older women than older men in the general population. Furthermore, the incidence of aneurysmal SAH rises with age, especially in women, possibly related to estrogen hormone differences in women and men and changes that result after menopause affecting aneurysm development.^23,28^

While rates for women showed a general decrease in mortality, men saw a marginal increase, particularly from 2014 to 2020. This may be indicative of a complex interplay between physiological differences, healthcare access, and perhaps even biases in clinical management strategies between sexes. It has been shown that men are at greater risk of ntSAH than women, but in age groups greater than 75, women are at greater risk than men. It is crucial to examine these differential trends to develop sex-specific preventive and therapeutic interventions.

With respect to racial and ethnic stratification, all groups exhibited a decrease in AAMR, albeit to varying extents. Xia’s article, which spanned between 2007 and 2017, showed an unchanged incidence of spontaneous SAH in Asian, Hispanic, and non-Hispanic whites but an increased incidence in Black patients. This possibly suggests that either incidence has particularly increased in the Asian cohort since 2017, or that there is greater mortality in this cohort after hemorrhage recently.

The COVID-19 epidemic posed several new health challenges since 2017, specifically affecting a greater number of non-White patients. It is possible that COVID-19, race, and ethnic differences played a role in this mortality. COVID-19 is implicated in vasculitis of small and medium-sized arteries, which can lead to endothelial inflammation of various vascular beds and cause non-aneurysmal ntSAH. The multiple organ dysfunction seen in patients with SAH and COVID-19 was far more pronounced than in patients with COVID-19 without SAH. Patients with COVID-19 who had SAH had higher mortality than patients without SAH.^29^ COVID-19 affected the management of ntSAH patients with fewer COVID-19 patients likely to undergo surgery, especially in elderly populations, and implementation of proning and medications that would otherwise complicate ntSAH management. Although unattainable by a query from the CDC WONDER database, a nuanced approach is needed to unravel the contributing socio-economic, genetic, and healthcare factors in these disparate outcomes.

The findings in this study illuminate geographical discrepancies related to urbanization. While all urbanization groups witnessed a decrease in mortality rates from 1999 to 2020, micropolitan areas remained persistently higher, and along with noncore areas stabilized instead of decreasing. Regional variances further complicate the landscape of ntSAH mortality, with the Western US exhibiting significantly higher mortality. While there are several academic centers in the West, the geographic distance between centers is greater than those of the Eastern and Central US, which could have affected transit times for patients transported to academic centers in the West. Interestingly, significant differences were noted at the state level, highlighting the influence of local healthcare infrastructure, policy, and possibly environmental factors.

Regional and urban differences in mortality may reflect different levels of access to healthcare resources. Micropolitan areas have a greater incidence of vascular risk factors, such as cigarette smoking, diabetes, and hypertension, which have been associated with greater predisposition to, and worse outcomes following ntSAH. Mariajoseph et al contend that this may explain the higher rates of mechanical ventilation seen in non-metropolitan location transfers after aneurysmal SAH, which yields greater rates of rebleeding due to delays in transfer.^30^ Furthermore, ntSAH care requires unique expertise, which has led to greater regionalization among higher-volume centers. Changes in mortality may suggest delayed transport to comprehensive stroke centers, which perform highly specialized endovascular and surgical procedures and have neuro-intensive care units. Care at high-volume teaching hospitals is more likely to reduce mortality and readmissions with multidisciplinary team-based interventions involving neurointensivists, neurosurgeons, endovascular specialists, nursing care, social work, and rehabilitation services. Older patients with ntSAH may require prompt consideration for referral to high-volume teaching hospitals to reduce this mortality.

## Limitations

There are inherent limitations to this study that warrant consideration. The employment of the ICD codes and the dependency on death certificates for data collection may engender some level of misclassification of ntSAH as a cause of death. Additionally, advancement in diagnostic capabilities and the increasing use of electronic health records may contribute to changes in the recording of ntSAH on death certificates, potentially influencing trends without necessarily reflecting actual changes in ntSAH-related mortality rates among the older population. The CDC WONDER database from which this study draws its data lacks granularity on disease characteristics such as clinical symptoms, laboratory markers, imaging studies, or other prognostic indicators. These limitations collectively circumscribe the ability to draw definitive conclusions and suggest that the findings should be interpreted cautiously.

## Conclusions

The study reveals that after an initial period of decline, the AAMR for ntSAH-related mortality among older adults has discernibly increased from 2013 to 2020 in the United States. This rise is nuanced by demographic variables such as sex, race and ethnicity, and geographic location, necessitating a multi-dimensional approach for its interpretation and management. Targeted health policy measures are imperative to address the escalating burden of ntSAH in the older population, with an emphasis on refined risk stratification, timely intervention, and the mitigation of persistent disparities.

## Data Availability

Data is available publicly

https://wonder.cdc.gov/mcd-icd10.html

## Acknowledgments

No acknowledgements

